# Border Restriction as a Public Health Measure to Limit Outbreak of Coronavirus Disease 2019 (COVID-19)

**DOI:** 10.1101/2020.10.29.20222190

**Authors:** Kwok Wang Chun, Wong Chun Ka, Ma Ting Fung, Ho Ka Wai, Fan Wai Tong Louis, Chan King Pui Florence, Chan Shung Kay Samuel, Tam Chi Chun Terence, Ho Pak Leung

**Affiliations:** Department of Medicine, Queen Mary Hospital, Hong Kong SAR, China; Department of Statistics, University of Wisconsin, Madison, United States of America; Department of Astronomy, University of Wisconsin, Madison, United States of America; Department of Mathematics, Indiana University, Bloomington, United States of America; Department of Microbiology, University of Hong Kong, Hong Kong SAR, China

## Abstract

**Background:** Coronavirus Disease 2019 (COVID-19) led to pandemic that affected almost all countries in the world. Many countries have implemented border restriction as a public health measure to limit local outbreak. However, there is inadequate scientific data to support such a practice, especially in the presence of an established local transmission of the disease.

**Method:** A novel metapopulation Susceptible-Exposed-Infectious-Recovered (SEIR) model with inspected migration was applied to investigate the effect of border restriction between Hong Kong and mainland China on the epidemiological characteristics of COVID-19 in Hong Kong. Isolation facilities occupancy was also studied.

**Results:** At *R*_0_ of 2·2, the cumulative COVID-19 cases in Hong Kong can be reduced by 13·99% (from 29,163 to 25,084) with complete border closure. At an in-patient mortality of 1·4%, the number of deaths can be reduced from 408 to 351 (57 lives saved). However, border closure alone was insufficient to prevent full occupancy of isolation facilities in Hong Kong; effective public health measures to reduce local *R*_0_ to below 1·6 was necessary.

**Conclusion:** As a public health measure to tackle COVID-19, border restriction is effective in reducing cumulative cases and mortality.

**Article summary:** *Strengths and limitations of this study:*

- A novel metapopulation SEIR model with inspected migration was developed to investigate the epidemiological characteristics of COVID-19 in Hong Kong, Guangdong and the rest of China (excluding Hubei) in the presence or absence of border restriction.
- The presented model is also suitable for further analysis of other emerging infectious diseases.
- Border restriction is an effective public health measure in reducing cumulative cases and mortality for COVID-19.
- Interaction was assumed to be well-mixed within patch. The spatial effect in disease transmission within each patch is ignored, which can have a non-trivial effect on the dynamic of infectious disease.
- The proposed model is deterministic in nature which ignores the randomness in migration and in the interactions among people; a stochastic model would be more realistic especially early in the disease.

## Introduction

In December 2019, there was an outbreak of Coronavirus Disease 2019 (COVID-19) (COVID-19) in Wuhan, China (1). As the amino acid sequences of the seven replicase domains used for classification in this coronavirus species were found to be 94·6% identical with SARS-CoV (2), the International Committee on Taxonomy of Viruses designated this novel virus as Severe Acute Respiratory Syndrome CoronaVirus 2 (SARS-CoV-2). Phylogenetic analysis of the full-length viral genome, RNA polymerase and S gene sequences showed a highest identity to a BatCoV RaTG13 previously detected in an intermediate horseshow bat (*Rhinolophus affinis*) from Yunnan Province, providing evidence for a bat origin of SARS-CoV-2. It was shown that 55% of cases with onset before 1^st^ January 2020 were linked to Huanan Seafood Wholesale Market in Wuhan, whereas later cases were predominantly mediated by human-to-human transmission (3). Transmission through asymptomatic contact also seemed highly probable (4), a feature that was not previously seen with SARS-CoV or MERS-CoV.

Clinical spectrum of SARS-CoV-2 infection ranges from flu-like illness to pneumonia with rapid progression to acute respiratory distress syndrome (ARDS) and death (1, 5-7). Among hospitalized patients, 32% to 51% had underlying disease and 26% to 32% of them required intensive care unit admission. The fatality rates for hospitalized COVID-19 patients varies between 0·6% to 15% (1, 6, 7) but the true disease-specific mortality rate is unclear because the proportion of asymptomatic and mild infections remains uncertain.

COVID-19 rapidly evolved and became a pandemic. As of 20^th^ July 2020, there were more than 14 million COVID-19 over the world (8). To limit the scale of local disease outbreak, many countries implemented travel restriction towards travellers from regions with severe COVID-19 outbreak and even all other countries, despite the World Health Organization (WHO) advising against implementing travel restriction as a public health measure to tackle COVID-19.

Hong Kong is a Special Administrative Region of the People’s Republic of China and border control exists between the two regions. Owing to the tight geographical and socio-economic ties, more than forty-million individuals travelled from mainland China to Hong Kong in a year (9). China was the earliest country with COVID-19 outbreak. On 23^rd^ January 2020, Hong Kong confirmed its first imported case of COVID-19 from Hubei (10). In the subsequent weeks, the number of imported cases rapidly rose despite initiation of various public health measures. Medical professionals and the general public repeatedly urged the Hong Kong government to close the Hong Kong-Chinese border to stop further influx. However, some questioned the effectiveness of such measure as there was already sign of local transmission in Hong Kong. Some believed that border restriction is not useful in the presence of established local transmissions as the final disease burden might be primarily driven by local transmission instead of importing of foreign cases.

To date, there is inadequate scientific data to support border restriction as a public health measure to limit local outbreak of an emerging infectious disease in the presence of an established local transmission. The objective of this study is to assess the impact of border restriction on cumulative caseload and hospital occupancy with a novel metapopulation Susceptible-Exposed-Infectious-Recovered (SEIR) model with inspected migration. Projection of COVID-19 epidemiology in Hong Kong and mainland China will be performed as an illustration.

## Method

In this study, a novel metapopulation SEIR model with inspected migration was applied to investigate the epidemiological characteristics of COVID-19 in Hong Kong, Guangdong and the rest of China (excluding Hubei) in the presence or absence of border restriction. Guangdong was separately analyzed from the rest of China because Guangdong province had significantly higher confirmed cases per population (11·7 per million) than the rest of China (excluding Hubei) (9·5 per million) as of 20^th^ February 2020. Hubei province, with the highest case density in China (1048·4 per million), was excluded from analysis as all Hubei-Hong Kong travel was banned after the Wuhan lockdown on 23^rd^ January 2020. Real world data up to 8^th^ February 2020 was used.

### Metapopulation SEIR Model with Migration

SEIR type models are commonly adopted to simulate epidemiology of infectious disease of a single region over time. It is based on a system of ordinary differential equations (ODE) that governs the number of 4 types of individuals: susceptible (S), exposed but latent (E), infectious (I), and recovered (or death) (R). Conventional single-patch SEIR models are not suitable for studying the impact of border restriction of an emerging infectious disease. A novel modified metapopulation SEIR model with inspected migration was used in this study. In addition to simulating population migration, parameters such as efficiency of custom inspection in blocking infected travellers were also being incorporated. Details of the model were described in Appendix 1.

### Assumption

It was assumed that there were no vital dynamics and well-mixed within patch for simplicity. Disease transmission between patches was assumed to be contributed by migration only and not by other means such as animal or environmental vectors. Since the rest of the world had negligible impact on the projection in the model at time of simulation, the interaction between Hong Kong, mainland China and the rest of the world was ignored for a very reasonable simplification. Reinfection was assumed to be not possible.

### Real life epidemiological data

The population sizes of Hong Kong, Guangdong and the rest of China (excluding Hubei) at the time of analysis were 75,241,000, 113,460,000 and 1,222,750,000 (11) respectively. As of 7^th^ February 2020, there were 26, 1034 and 5787 cases of laboratory confirmed COVID-19 patients in the three region respectively according to Hong Kong Department of Health and China Centre for Disease Control (CDC) data. (12)

### Model Parameters

The mean incubation and infectious period was taken as 5·2 and 5·0 days respectively (3). Coronavirus transmissibility has been hypothesized to reduce as temperature rises (13), hence *R*_0_ is set to be inversely correlated with temperature. *R*_0_ was set to linearly reduce from initial value at 18·0°C to 0 at 25·0°C. The temperature threshold was set by referencing Hong Kong temperature in the summer of 2003 when SARS, which was also caused by coronavirus, subsided. Temperature in the projected period was modelled based on 2019 data released by the Hong Kong Observatory (14). To explore the effect of border crossing restriction, we conducted simulations with 200,000 and 0 individuals travelling from mainland China to Hong Kong per day. We assumed 70% were from Guangdong and 30% were from the rest of China (excluding Hubei), based on the previous data from Hong Kong Immigration Department (15). Efficiency of Immigration Department in blocking visitors in latent period (1 − *σ*) was taken as 50% by assuming household close contact of infected individuals were all quarantined and non-household close contact were not quarantined. Efficiency of Immigration Department in blocking visitors in infectious period (1 − *θ*) was taken as 99% by assuming that body temperature monitoring and compulsory health declaration at the Immigration Department were 99% efficient. The listed model parameter is summarized in Table 1. Simulation with multiple initial *R*_0_ values were performed, starting from 2.2, down to Re 1·6 at 0·1 intervals.

### Isolation facility occupancy

The Hong Kong public health system had a maximum of 952 isolation beds in 490 isolation single rooms according to the data from Hospital Authority press conference on 1^st^ March 2020. It was assumed that all isolation facilities were used exclusively for COVID-19 purposes.

## Results

### Effect of complete border closure on case number and mortality

We applied the novel metapopulation SEIR model with inspected migration to project the case number in the presence or absence of complete border closure. At *R*_0_ of 2·2, reduction in number of daily travellers from 200,000 to 0 starting 8^th^ February 2020 would decrease the cumulative COVID-19 cases in Hong Kong by 13·99% from 29,163 to 25,084. At an in-patient mortality of 1·4%(16), the number of deaths can be reduced from 408 to 351 (57 lives saved). At *R*_0_ of 1·6 – 2·1, complete border closure was projected to cause a 11·54 – 13·71% reduction in cumulative cases and mortality (Figure 1 and Table 2). The results suggested that even in the presence of established local transmission, travel restriction remains an effective measure to reduce the cumulative cases in the recipient region. COVID-19 associated mortality can also be decreased with this measure.

**Figure 1.**
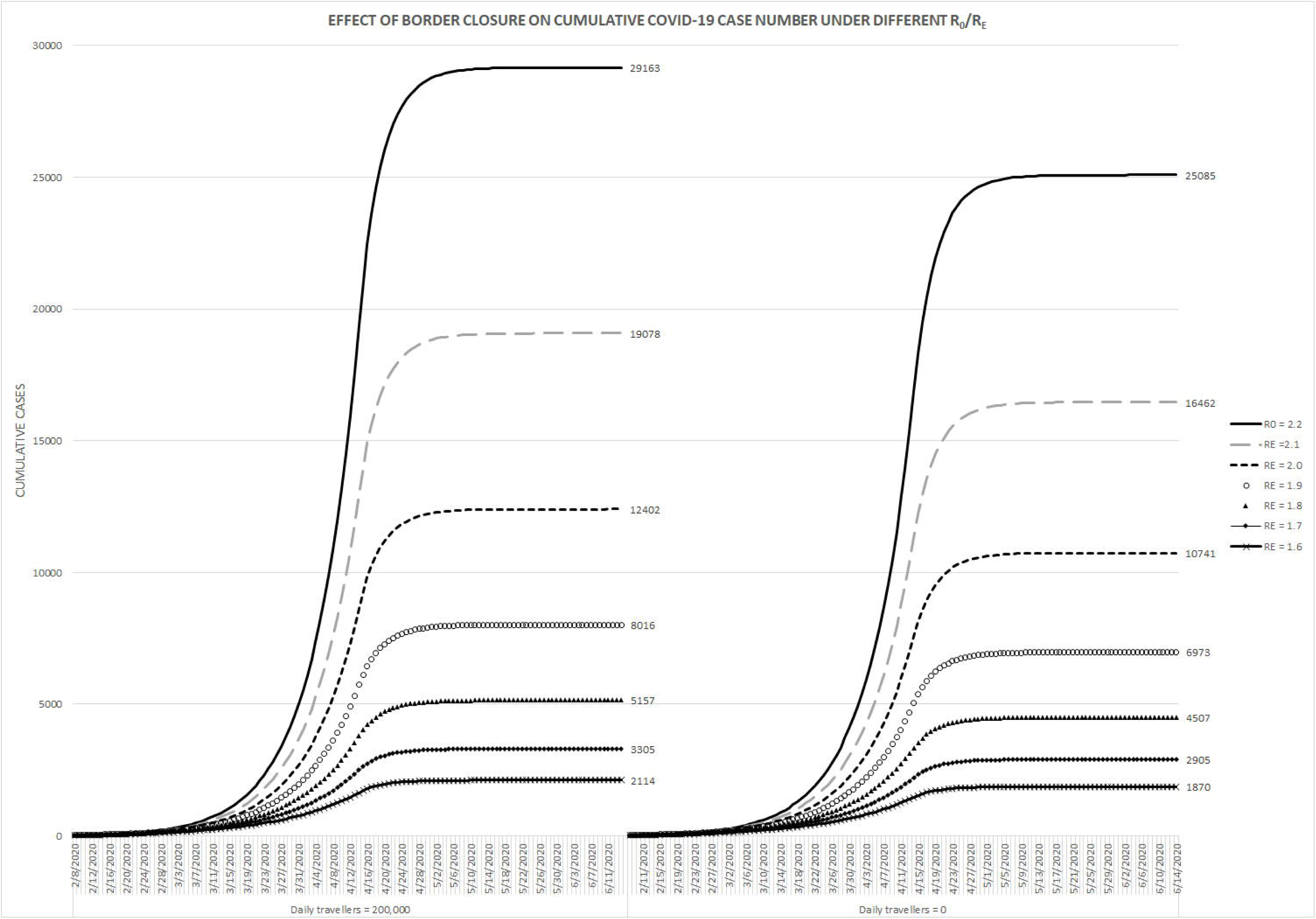
Effect of complete border closure on the projected cumulative case number of COVID-19 across different reproductive number over time

### Effect of public health measures on projected isolation facility demand

Local *R*_0_ of an infectious disease is partially dependent on effectiveness of public health measures implemented in a region (18). It can be in the form of contract tracing and quarantine system, or social distancing policies such as school cessation. For Hong Kong, at *R*_0_ of 2·2, the projected number of concurrent isolation facilities required to accommodate all infected individuals at the peak of the epidemic is 5,782 even with border restriction; this translates into the additional need of 5,292 new isolation rooms. Maintaining complete border closure and having effective public health measures to keep *R*_0_ below 1.6 is required to allow Hong Kong to meet its isolation room requirement. Other permutations are shown in Table 3, and graphically represented in Figure 2.

**Figure 2.**
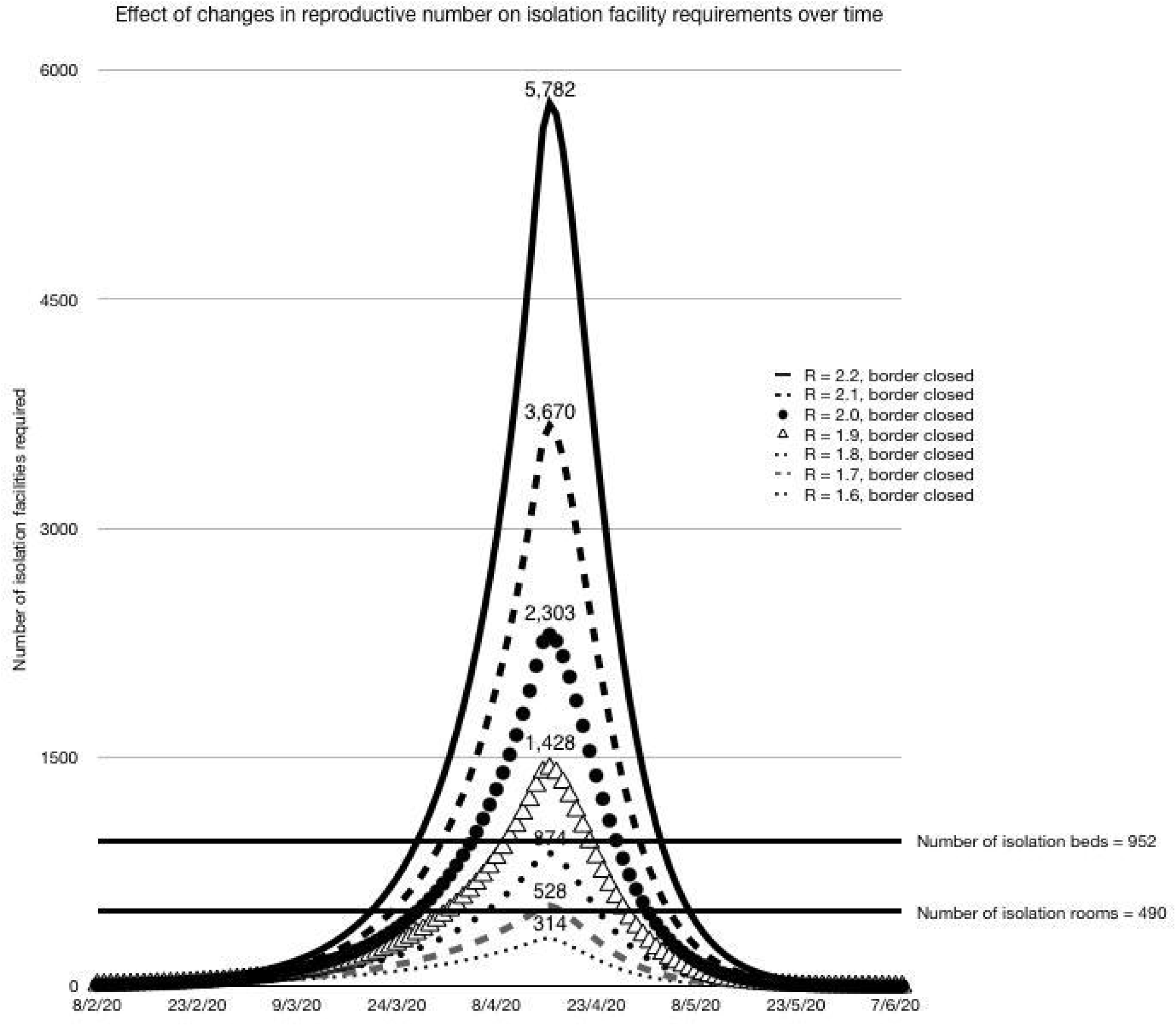
Effect of changes in reproductive number on isolation facility requirements over time

## Discussion

Countries or cities with a high population density and aged population including Hong Kong is at risk of severe outbreak of emerging infectious diseases such as COVID-19. As the disease is spreading rapidly in multiple continents, many countries implemented border restrictions towards regions with severe outbreak in order to reduce local case number and mortality. This is particularly important for developing countries with inadequate medical resources to tackle massive local outbreak. However, the WHO advised against utilizing travel restriction as an infection control measure. Furthermore, it is particularly challenging to implement border restriction in certain regions due to political, social and economical reasons. To date there is inadequate scientific data to support border restriction as a public health measure to limit the scale of local outbreak in the presence of an established local transmission. Using Hong Kong and mainland China as an example, we quantitatively illustrated border restriction is effective in reducing cumulative caseload, mortality and healthcare facility occupancy with a novel metapopulation SEIR model with inspected migration. It was projected that complete border closure will result in meaningful reduction of cumulative cases (4079 cases at *R*_0_ of 2·2), mortality (57 deaths at 1·4% in-patient mortality) and a delay in isolation facility overload in Hong Kong.

It is important to emphasize that in our projection, border closure alone is insufficient to prevent healthcare overload, as measured by isolation facilities occupancy. Effective and targeted public health intervention to slow local transmission and reduce local *R*_0_ is needed. It can be in the form of universal usage of surgical mask, contract tracing and quarantine system, or social distancing policies such as school cessation. The outbreak on Princess Diamond Cruise clearly illustrated the limitation in outbreak control by border restriction solely with no public health intervention. In early 2020, a number of passengers on Princess Diamond Cruise were found to be have COVID-19. Despite there was no further import of COVID-19 cases onto the cruise after the immediate quarantine, there was still rapid rise in the number of COVID-19 cases on the cruise. It was believed that insufficient on-board personal protective equipment and inadequate social distancing were the causes of the unfortunate event. To date, of the 3,711 individuals on the cruise, 624 of 3011-tested passengers were diagnosed with COVID-19 (16·7%). Unfortunately, implementation of strict public health measures may not be feasible to combat COVID-19 in many regions. For instance, social distancing may not be feasible due to environmental, economical, cultural or religion reasons, and there may be a shortage of trained personnel and facilities for performing contact tracing and quarantine.

In the past few months, multiple regions had exponential rise in COVID-19 cases which caused extreme stress to their local health care system. In Wuhan, which was the epicenter of the COVID-19 outbreak in China, severe shortage in isolation facilities urged urgent construction of multiple temporary hospitals. COVID-19 related mortality in regions with severe outbreak tend to be higher due to relative shortage of medical resources outweigh demand. Advanced life support facilities such as intensive care unit, ventilators, extracorporeal membrane oxygenation (ECMO) machines and anti-viral medications were essential in severe COVID-19 cases but their availability is limited. In addition, COVID-19 also severely hinder other non-COVID-19 related medical services. In Hong Kong, although the total confirmed COVID-19 cases is less than the available isolation facilities at the moment, a significant proportion of other less urgent medical services include elective investigations and surgeries have been suspended to reserve resources for COVID-19. In less resourceful regions, the effect may even be more pronounced. Although morbidity and mortality caused by such service suspension are not included in the official COVID-19 statistics, the effects cannot be overlooked. Furthermore, uncontrolled local epidemic can cause outbreaks in other regions with close ties and lead to a pandemic situation. The damage brought by a severe local outbreak of COVID-19 is unbearable. Therefore, it is paramount for governments around the world to prevent or limit scale of local outbreak. As suggested by our projection, border restriction against regions with severe outbreak could reduce local caseload, mortality and isolation facilities occupancy. Furthermore, aggressive and efficient public health measures to reduce local *R*_0_ is necessary.

### Strength of the model

The spread of infectious disease is closely related to the migration of population between regions (19, 20). Conventional single-patch SEIR models are not suitable for such analysis. A novel metapopulation SEIR model with inspected migration was specifically developed for this purpose. In addition to COVID-19, the developed model can be used to perform projection for other emerging infectious diseases in the future. Furthermore, parameters such as effectiveness of custom inspection were included to improve accuracy of projection. The presented model is also suitable for further analysis of other emerging infectious diseases.

### Limitation

Firstly, interaction was assumed to be well-mixed within patch. The spatial effect in disease transmission within each patch is not directly addressed in the model, which can have a non-trivial effect on the dynamic of infectious disease (21). Secondly, the proposed model is deterministic in nature which ignores the randomness in migration and in the interactions among people; a stochastic model would be more realistic especially early in the disease. Thirdly, key parameters such as rate of spread is still unclear so we assumed a parametric form of the rate of spread with reference to 2003-SARS. In general, parameter calibration can be performed by some criteria, for example, minimizing residuals sum of square between the historical and fitted infected cases. Meanwhile, missing information, such as travel history across regions, leads to crucial statistical uncertainty. A stochastic metapopulation migration model to explore the corresponding statistical properties with data would be a fruitful direction in the future. While the above shortcomings may be the expected tradeoff between computation time and model simplicity, it will not negate the signal that core message that border restriction reduces cumulative case, mortality and delay healthcare system exhaustion. Lastly, economic impact is beyond the scope of this study. While full border closure can have a negative impact on the economy, one cannot ignore the negative economic impact from an otherwise preventable major outbreak. At time of writing, COVID-19 was perceived to have developed into a pandemic situation and global stock market plummeted with The Dow Jones Index and Hang Seng Index both fell more than 9% percent since 2/1/2020.

## Conclusion

As a public health measure to tackle COVID-19, border restriction is effective in reducing cumulative cases and mortality. Hospital occupancy can be reduced but effective public health measures to achieve significant reduction in *R*_0_ would be necessary to prevent full occupancy of available isolation facilities.

## Data Availability

No data available

This research did not receive any specific grant from funding agencies in the public, commercial, or not-for-profit sectors.

No potential conflict of interest was reported by the authors.

The lead author (the manuscript’s guarantor) affirms that the manuscript is an honest, accurate, and transparent account of the study being reported; that no important aspects of the study have been omitted; and that any discrepancies from the study as planned (and, if relevant, registered) have been explained.

Ethical approval is not required as this study does not involve patients.

Our study is reported according to the GATHER statement. ·

## Data sharing

No additional data available.

## Table and Figures

Table 1. Model Parameters

Table 2. Effect of complete border closure on the projected cumulative COVID-19 case & mortality at *R*_0_ = 2 · 2 and different *R*_0_ down to 1·6

Table 3. Projected isolation facility deficit at *R*_0_ = 2 · 2 and different *R*_0_ down to 1·6 (Assuming complete border closure & 100% isolation / hospitalization rate)

