## Supplementary material for "Border Restriction as a Public Health Measure to Limit Outbreak of Coronavirus Disease 2019 (COVID-19)": Table 1

Table 1. Model Parameters

| Parameter | Value | Rationale/ Assumption |
| --- | --- | --- |
| Latent period | 5.2 days | As reprorted in Li et al. (3) |
| Infective period | 5.0 days | Time from symptom onset to establishing diagnosis, getting isolated and rendering effectively non-infectious in Hong Kong. |
| Initial maximal $R_{0}$ | 2.2 | As reported in Li et al.(3) |
| Temperature at which $R_{0}$reduce to 0 | 25.0 degree Celsius | Novel coronavirus transmissibility Hypothesized to reduce as temperature rises (19). Threshold set with reference to temperature in Hong Kong in 2003 when SARS subsided near summer. |
| Efficiency of Immigration Department in blocking visitors in latent period ($\sigma$) | 50% | Assumed household close contact of infected individuals are all quarantined and non-household close contact are not quarantined. |
| Efficiency of Immigration Department in blocking visitors in infectious period ($\theta$) | 99% | Assumed temperature monitoring and compulsory health declaration process at Immigration Department is 99% efficient. |
| In-patient mortality rate (lower bound) | 1.36% | As reported by Zhong et al. (17) |
| In-patient mortality rate (upper bound) | 4.3% | As reported by Wang et al. (1) |
