## Supplementary material for "Border Restriction as a Public Health Measure to Limit Outbreak of Coronavirus Disease 2019 (COVID-19)": Table 2

Table 2. Effect of complete border closure on the projected cumulative COVID-19 case & mortality at $R_{0}=2\cdot2$ and different $R_{0}$ down to 1·6

|  | Without border closure (Daily traveller 200,000) | Complete border closure (Daily traveller 0) | | | |
| --- | --- | --- | --- | --- | --- |
|  |  | Case reduction | Death averted | | Percent  reduction |
|  |  |  | (@4.30%) | (@1.36%) |  |
| $R_{0}$ 2.2 | 29,163 | 4079 | 175 | 56 | 13.99% |
| $R_{E}$ 2.1 | 19,078 | 2616 | 112 | 35 | 13.71% |
| $R_{E}$ 2.0 | 12,402 | 1661 | 71 | 23 | 13.39% |
| $R_{E}$ 1.9 | 8,061 | 1088 | 45 | 14 | 13.50% |
| $R_{E}$ 1.8 | 5,157 | 650 | 28 | 9 | 12.60% |
| $R_{E}$ 1.7 | 3,305 | 400 | 17 | 5 | 12.10% |
| $R_{E}$1.6 | 2,114 | 244 | 11 | 4 | 11.54% |
