## Supplementary material for "Border Restriction as a Public Health Measure to Limit Outbreak of Coronavirus Disease 2019 (COVID-19)": Table 3

Table 3. Projected isolation facility deficit at $R_{0}=2\cdot2$ and different $R_{0}$ down to 1·6 (Assuming complete border closure & 100% isolation / hospitalization rate)

|  | Maximum concurrent facility needed* | Additional isolation facilities required | | | |
| --- | --- | --- | --- | --- | --- |
|  |  | Single rooms | | Isolation beds | |
|  |  | Extra rooms needed | Date of reaching 100% occupancy | Extra beds needed | Date of reaching 100% occupancy |
| $R_{0}$ 2.2 | 5,782 | 5,292 | 21-Mar-2020 | 4,830 | 28-Mar-2020 |
| $R_{E}$ 2.1 | 3,670 | 3,180 | 24-Mar-2020 | 2,718 | 31-Mar-2020 |
| $R_{E}$ 2.0 | 2,303 | 1,813 | 28-Mar-2020 | 1,351 | 05-Apr-2020 |
| $R_{E}$ 1.9 | 1,428 | 938 | 01-Apr-2020 | 476 | 10-Apr-2020 |
| $R_{E}$ 1.8 | 874 | 384 | 07-Apr-2020 | N/A | N/A |
| $R_{E}$ 1.7 | 528 | 38 | 14-Apr-2020 | N/A | N/A |
| $R_{E}$ 1.6 | 314 | N/A | N/A | N/A | N/A |
| *^ Current capacity with 490 isolation rooms & 952 isolation beds* | | | | | |
