## Appendix 1 for "Border Restriction as a Public Health Measure to Limit Outbreak of Coronavirus Disease 2019 (COVID-19)"

In this study, a novel metapopulation SEIR model with inspected migration was applied.

To model migration across borders and assess its impact on the epidemiological spread, we let $m$ be the number of patches/sub-populations and consider an $m$-patch metapopulation SEIR model with migration. We denote $N_{i}$to be the effective population size of patch. Let $(S_{i},E_{i},I_{i},R_{i})$ be the number of susceptible, exposed but latent, infectious and recovered (or death) individuals respectively in the $i$-th patch, viewed as functions of time $t$.

With rate $\mu_{ij}(t)$ at time $t$, individuals in patch $i$ try to migrate to path $j$. Among the infected individuals $I_{i}$, only a fraction $\theta_{ij}\in[0,1]$ will pass through the border from $i$ to $j$ due to custom inspection (such as temperature monitoring and compulsory health declaration). The remaining $(1-\theta_{ij})$fraction will be sent back to patch $i$. In particular, if $\theta_{ij}=0$, then the inspection is doing a perfect job in blocking all infected individuals at the border. Likewise, among the exposed but latent individuals $E_{i}$, only a fraction $\sigma_{ij}\in[0,1]$ will pass through the border from $i$ to $j$ and the remaining $(1-\sigma_{ij})$fraction will be sent back to patch $i$. Note that different from traditional single-patch SEIR model, the sub-population (patch) size $N_{i}=S_{i}+E_{i}+I_{i}+R_{i}$ is no longer constant over time due to migration (traveling across borders).

The mathematical representation of the model is as follows: For $i=1,\ldots,m$, where $m$ is the total number of patches,

$$\frac{dS_{i}}{dt}=-M_{i}S_{i}+\sum_{j:j\neq i} \mu_{ji}S_{j}-\frac{\beta I_{i}S_{i}}{N_{i}}$$

$$\frac{dE_{i}}{dt}=-\left[ \sum_{j:j\neq i} \sigma_{ij}\mu_{ij} \right]E_{i}+\sum_{j:j\neq i} \sigma_{ji}\mu_{ji}E_{j}-\alpha E_{i}+\frac{\beta I_{i}S_{i}}{N_{i}}$$

$$\frac{dI_{i}}{dt}=-\left[ \sum_{j:j\neq i} \theta_{ij}\mu_{ij} \right]I_{i}+\sum_{j:j\neq i} \theta_{ji}\mu_{ji}I_{j}-\gamma I_{i}+\alpha E_{i}$$

$$\frac{dR_{i}}{dt}=-M_{i}R_{i}+\sum_{j:j\neq i} \mu_{ji}R_{j}+\gamma I_{i}$$

where $N_{i}=S_{i}+E_{i}+I_{i}+R_{i}$ is the population size of path $i$**.**

Interpretation of the parameters as follows:

1. $\mu_{ji}$ is the migration rate from patch $j$ to patch $i$, and we set $\mu_{ii}=0$ as individuals that are staying in the same patch $i$. A partly closed border model can be obtained by setting some $\mu_{ji}$ equals zeros.
2. $M_{i}=\sum_{j:j\neq i} \mu_{ij}$is the total migration rate from the patch $i$.
3. ${\sigma_{ji,}\theta}_{ji}\in[0,1]$are discount factors taking into account ability of custom inspection to block exposed but latent and infectious individuals respectively across the border from patch $j$ to patch $i$, and we set $\theta_{ii},\sigma_{ii}=0$ as individuals are staying in the same patch $i$. A perfect border screening model can be obtained by setting $\theta_{ji},\sigma_{ji}$ to zeros.
4. $N_{i}=N_{i}(t)\in(0,\infty)$is the effective population size of patch $i$. Note that we can keep $N_{i}$ fixed at the initial sub-population (patch) size $N_{i}(0)$. It is reasonable especially when the net migrant of each patch is closed to zero.
5. $\alpha\in(0,1)$ is the incubation rate, which controls the rate of exposed but latent individuals becoming infectious, which is equals to inverse of latent period.
6. $\beta\in(0,\infty)$ is the infectious rate, which controls the rate of transmission of the disease between a susceptible and an infectious individual, which equals to $R_{0}\gamma$. It is possible to introduce $\beta$ that varies with time and other variables, such as climatological variables.
7. $\gamma\in(0,1)$ is the rate at which infectious individuals are rendered non-infectious, including recovery, death and getting quarantined. It is equal to the inverse of the infectious period.
8. Cumulative cases are the sum of locally infected individuals and imported infected individuals.
